# All-cause and cardiac mortality in relation to smoke exposure nine years after the Hazelwood coal mine fire

**DOI:** 10.1101/2025.01.14.25320562

**Authors:** Karen Walker-Bone, Caroline X. Gao, Catherine L Smith, Tingting Ye, David Brown, Natasha Kinsman, Jillian F Ikin, Matthew Carroll, Michael J Abramson, Yuming Guo, Tyler J Lane

## Abstract

**Background:** In 2014, a coal mine fire in rural Australia shrouded nearby towns in smoke for several weeks. In response to community concerns and a lack of available evidence, the Victorian Department of Health commissioned a long-term health study. This paper evaluates smoke effects on mortality 9-years after the fire.

**Methods:** In 2016-17, 4,056 members of the exposed community and a demographically-similar control were recruited into a cohort study. A third of these (2,725 (67%)) consented to linkage of their data. To estimate individual-level smoke exposure, participant’s time-location diaries were blended with estimates of hourly fire-related PM_2.5_ based on meteorological and chemical transport models. All-cause mortality data were obtained up to mid-2023 by linkage with the National Death Index. Cardiac-related deaths were identifiable up to December 2021 and predicted thereafter to mid-2023. Differences in all-cause mortality were evaluated with Cox proportional-hazards models and differences in cardiac mortality with a competing risk survival model.

**Results:** Across 6-7 years of follow-up, 303 (11%) of the sample died. All-cause mortality was not associated with PM_2.5_ exposure. A 10µg/m^3^ increase in PM_2.5_ was associated with a 17% (95%CI: 1.01-1.36) higher risk of mortality from cardiac causes.

**Discussion:** PM_2.5_ from the mine fire was associated with elevated cardiac mortality between 2-9 years post-exposure. This finding concords with a growing body of evidence after acute exposure to wildfires and chronic exposures from household or ambient air pollution. Measures are needed to protect individuals from exposure to smoke during and after large fire events.

## 1 Introduction

During severe fire weather in mid-Summer 2014, embers from a grass fire in regional Victoria, Australia ignited the open-cut brown coal mine adjacent to the Hazelwood power station. The resultant coal mine fire burned for six weeks, enveloping nearby communities in dense smoke and ash. More than 7000 firefighters were deployed, 500 or so at a time, to suppress the fire and many firefighters were treated for smoke inhalation. At the same time, there were complaints from people living nearby of numerous symptoms, including headaches, blurred vision, cough, shortness of breath, epistaxis, fatigue, gastrointestinal symptoms (including nausea, vomiting and diarrhoea), and chest pain. Such were the levels of concern in the local community that there were two judicial Inquiries and calls for a research study. Ultimately it was decided that a longitudinal health study lasting at least 20 years should be performed to determine the long-term health effects of smoke. Accordingly, the Victorian Department of Health issued a request for tender for what would become the Hazelwood Health Study, which started later the same year (1).

Coal mine fires can occur wherever coal is found and many are currently burning globally, particularly where there are rich subterranean coal deposits, such as in China, India and the USA (2–4). Coal mine fires release polluting emissions including high concentrations of toxic gases, volatile organic compounds and trace elements including carbon monoxide, polycyclic aromatic hydrocarbons, benzene and particulate matter (PM). These include PM with a median aerodynamic diameter less than 10 μm (PM_10_) and fine PM less than 2.5 μm (PM_2.5_), which are fine enough to reach the alveoli. Some health effects of these emissions have received attention through studies of ambient urban background air pollution by PM_2.5_ and after exposure to humans during forest and/or peat fires or through studies after domestic coal use. However the effects of coal mine fire exposure on long-term health outcomes have received considerably less attention (2). One particular concern is mortality, given that several constituents of coal fire smoke have been found to be associated with increased mortality, including PM_2.5_ (all-cause mortality), carbon monoxide, benzene, formaldehyde, phenols and naphthalene (cancer), and some trace elements. Moreover, increased mortality rates have been shown after forest fire exposure by some (5–11), but not all (12,13) studies.

Our prior analysis of mortality found that coal mine fire smoke PM_2.5_ exposure was associated with an increase in injury-related deaths during the mine fire and cardiac-related deaths in the six months after the fire (14). However, this was an ecological time series study, with mine fire smoke exposure estimated based on residential areas recorded in the mortality database. These analyses could not account for individual-level smoke exposure as well as relevant and important individual-level confounding factors such as pre-fire health status, tobacco use and socio-economic deprivation.

In this paper, we build on our previous analyses of the mine fire effects on population-level mortality (14) using individual-level data collected from participants in the Hazelwood Health Study Adult Cohort (15). Survey data from the cohort were linked to the Australian National Death Index (NDI), which we analysed to answer the following research questions: 1. Was there any long-term impact of the mine fire on all-cause mortality or cardiac mortality after 6-7 years of follow-up? 2. Were individual levels of exposure to fire-related PM_2.5_ associated with all-cause mortality and/or cardiac mortality?

## 2 Methods

### 2.1 Study cohort

The Hazelwood Health Study Adult Cohort comprised people who completed the Adult Survey between May 2016 and February 2017. The electoral roll maintained by the Victorian Electoral Commission was chosen to sample people who were residents in two towns in eastern Victoria (electoral registration is compulsory for Australian citizens aged ≥18 years). The first town was Morwell, adjacent to the mine fire and most severely affected by smoke exposure (exposed group). A comparison (minimally or not exposed) group was recruited from people who, during the mine fire, were living in 16 selected statistical areas (SA1s) of Sale, a town approximately 65km away. These SA1s were chosen to reflect areas of Sale with characteristics as similar as possible to those of Morwell in terms of median age, household size, socioeconomic factors and population stability. Data from air pollution modelling showed that Sale had minimal smoke exposure during the mine fire (16). Residents identified as living in these locations were sent postal invitations to participate along with information leaflets about the study. In addition to completing the Adult Survey, participants were invited to provide written informed consent to link their data with NDI. More information about the Adult Cohort is available in our cohort profile (15).

### 2.2 Exposures

Air quality monitoring in Morwell and its surrounds during the mine fire were inadequate, particularly in the earliest days when smoke levels were at their most intense (1). Consequently, high resolution spatial and temporal estimates of hourly fire-related PM_2.5_ distribution were generated through emission, chemical transport and meteorological models (17). To determine Individual-level smoke exposure, Adult Survey participants provided time-location diaries for the mine fire period in 12- hour periods (15), and the diary data were then combined with the modelling output (16), see previous analyses of this cohort for more detail (15,17).

### 2.3 Outcome

Mortality was measured in survival days by linkage to the National Death Index (NDI) (18). The NDI captures all deaths occurring in Australia from 1980 onwards and is housed by the Australian Institute for Health and Welfare. Subject to ethical review, processes are in place for researchers to obtain access to the NDI for research purposes via record linkage for all deaths. Cohort data were linked to NDI-identified deaths up to June 2023. The underlying cause of death was available from the NDI up to, and including, deaths until December 2021 and could be used to identify cardiac-related mortality (ICD-10: I00-I99, G45, G46).

### 2.4 Risk Factors

The following risk factors were explored in the analyses (see Statistical Analysis below), all measured at the time of the survey: age, sex, educational attainment (“secondary up to year 10”, “secondary years 11-12”, “certificate/diploma/tertiary degree”), study site (Morwell or Sale), tobacco use (smoker status: “current”, “former with at least 100 lifetime cigarettes”, and “never” as well as tobacco pack-years), and self-reported pre-fire comorbidities (either self-reported angina, myocardial infarction, heart failure, stroke, COPD, cancer or diabetes). To control for socioeconomic differences between participants, we also included the Index of Relative Socioeconomic Advantage and Disadvantage (IRSAD) score for each participant’s Statistical Area Level 1 (SA1) based on 2016 census data (19).

### 2.5 Statistical analysis

To identify potential cardiac-related mortality in 2022-23 when the cause of death was not available, a predictive model was developed using a nested cross-validated (outer Loop: 5-fold and inner loop: 10-fold) XGBoost algorithm (20). This prediction model took advantage of the data-linkage study design and obtained potential predictors from the Adult Survey (e.g., demographics, smoking, alcohol consumption, socioeconomic status, psychological distress, and self-reported doctor-diagnosed conditions). The model included predictors extracted from three linked healthcare records: the Victorian Admitted Episodes Dataset (VAED) for hospital admissions (21), the Victorian Emergency Minimum Dataset for emergency presentations (22), and Ambulance Victoria electronic patient care records for ambulance attendances up to 30 June 2022 (23). These predictors included the total number of healthcare services utilised, as well as service utilisation for specific condition groups (classified by ICD-10 chapters and corresponding ambulance diagnostic groups), within the 5 years preceding the mortality event. Due to the high dimensionality of the data, we first ran the prediction model to identify the top 20 predictors and re-trained it with selected predictors to improve model accuracy.

Descriptive statistics were used to characterise the cohort. Differences in all-cause based on fire-related PM_2.5_ exposure were evaluated with Cox proportional-hazards models (24), while differences in cardiac-related mortality based on fire-related PM_2.5_exposure were evaluated with a competing risk survival model, which accounted for deaths due to other causes (25).

Survival was measured from the date of the Adult Survey until the end of follow-up linkage with NDI data (27 June 2023). Analyses were adjusted for risk factors that were added to an initial crude model in the following order: age, sex, educational attainment, IRSAD score, study site, tobacco use and comorbidities. To determine whether any risk factors influenced the relationship between exposure and outcome, we performed a series of moderator analyses, incorporating interactions between exposure and the moderator, using covariates from the fully-adjusted model. To enhance interpretability, continuous moderators were standardised into z-scores and the daily mean fire-related PM_2.5_ across the mine fire period was expressed in units of 10 µg/m³ and centred at 10 µg/m³.

Missing data were addressed with random forest multiple imputations (26) and pooled according to Rubin’s rules (27). The number of imputations was equivalent to the proportion of cases with missing data (10 imputed datasets). All analyses were conducted in R (28) with RStudio (29). Cleaning and analytical code are available on our public repository (30).

### 2.6 Ethics

This study was approved by the Monash University Human Research Ethics Committee as part of the Hazelwood Adult Survey and Health Record Linkage Study (Project ID: 25680; previously CF15/872 - 2015000389 and 6066). Participants provided written informed consent to linkage.

## 3 Results

### 3.1 Description

From 4,056 Adult Survey participants, 2,115 from Morwell (68%) and 610 (64%) from Sale consented to the linkage of their Survey responses with NDI data. Participants from both towns had similar age and sex distributions and similar levels of cigarette smoking exposure (pack years) at the time of the Survey. However, Sale participants had slightly higher levels of educational attainment and were more socio-economically advantaged than those from Morwell (Table 1). As expected, Morwell residents had much higher estimated PM_2.5_ exposure during the mine fire. In total, during the 6-7 years of follow-up, 234 deaths occurred among residents of Morwell and 69 among residents of Sale, equating to 11% of each group. While the proportion of cardiac-related deaths was higher in Morwell, it was not statistically significantly so.

**Table 1.** Descriptive statistics by study site.

|  | Morwell<br>*n* = 2,115 | Sale<br>*n* = 610 | *p*-<br>value |
| --- | --- | --- | --- |
| All-cause mortality | 234 (11%) | 69 (11%) | 0.884 |
| Cause of death* |  |  | 0.160 |
| Cardiac | 75 (3.5%) | 14 (2.3%) |  |
| *Actual* | 55 (2.6%) | 11 (1.8%) |  |
| *Predicted* | 20 (0.9%) | 3 (0.5%) |  |
| Other | 158 (7.5%) | 56 (9.2%) |  |
| *Actual* | 106 (5.0%) | 42 (6.9%) |  |
| *Predicted* | 53 (2.5%) | 13 (2.1%) |  |
| Survived | 1,881 (89%) | 541 (89%) |  |
| Daily mean exposure to fire-related PM<sub>2.5</sub> (µg/m<sup>3</sup>) | 11 (7-19) | 0 (0-0) |  |
| Age at survey | 60 [IQR: 48-70] | 60 [IQR: 47-72] | 0.543 |
| Male | 980 (46%) | 265 (43%) | 0.213 |
| Educational attainment |  |  | <0.001 |
| Secondary to year 10 | 671 (32%) | 139 (23%) |  |
| Secondary to year 12 | 421 (20%) | 99 (16%) |  |
| Certificate/diploma/tertiary degree | 1,011 (48%) | 369 (61%) |  |
| Missing | 12 | 3 |  |
| IRSAD scores | 833 [IQR: 782-889] | 898 [IQR: 844-952] | <0.001 |
| Missing | 57 | 0 |  |
| Smoking status |  |  | 0.013 |
| Non-smoker | 974 (46%) | 318 (53%) |  |
| Former smoker | 763 (36%) | 205 (34%) |  |
| Current smoker | 365 (17%) | 82 (14%) |  |
| Missing | 13 | 5 |  |
| Cigarette pack years (current/former smokers) | 15 [IQR: 5-30] | 15 [IQR: 6-30] | 0.455 |
| Missing | 21 | 4 |  |
| Any pre-fire comorbidity | 629 (30%) | 170 (28%) | 0.391 |

Notes: Continuous data presented by median and inter-quartile range (compared with Kruskal-Wallis tests); dichotomous/categorical data presented by number and percent (compared with Fisher's exact tests); \* cause of death predicted for *n* = 89 who had died in the most recent 18
months for which death notification was available but cause was not; p-value is based on comparison of combined predicted and actual causes of death.

### 3.2 Prediction model

The prediction model with a total of 225 observations, including 67 known cardiac deaths as training data, had a cross-validation Area Under the Curve of 0.76. With a probability cut-off of 0.45, the algorithm had a sensitivity of 0.98 and specificity of 0.80 on the training data. The model predicted an additional 23 cardiac deaths among 93 new mortality records that did not specify a cause of death. The variable importance from the final prediction model is provided in Figure S1, which indicates mine fire exposure is one of the key predictors differentiating between cardiac and non-cardiac deaths.

### 3.3 Main results

Figure 1 and Table S1 summarise the results of the Cox proportional hazards (all-cause mortality) and competing risks (cardiac mortality) survival models, providing estimated Hazard Ratios (HRs) and 95% confidence intervals for each model. Model 1 for each outcome shows the crude association/effect of PM_2.5_ with overall mortality and cardiac mortality respectively. Models 2 to 6 show the effect of PM_2.5_ after adjusting for confounders which are added incrementally into the model. In the plots below, Model 6, each confounder was assessed for effect modification by adding the interaction between the potential effect modifier and PM_2.5_ to Model 6, with only the resulting interaction and their main effects shown in the plot.

**Figure 1.**
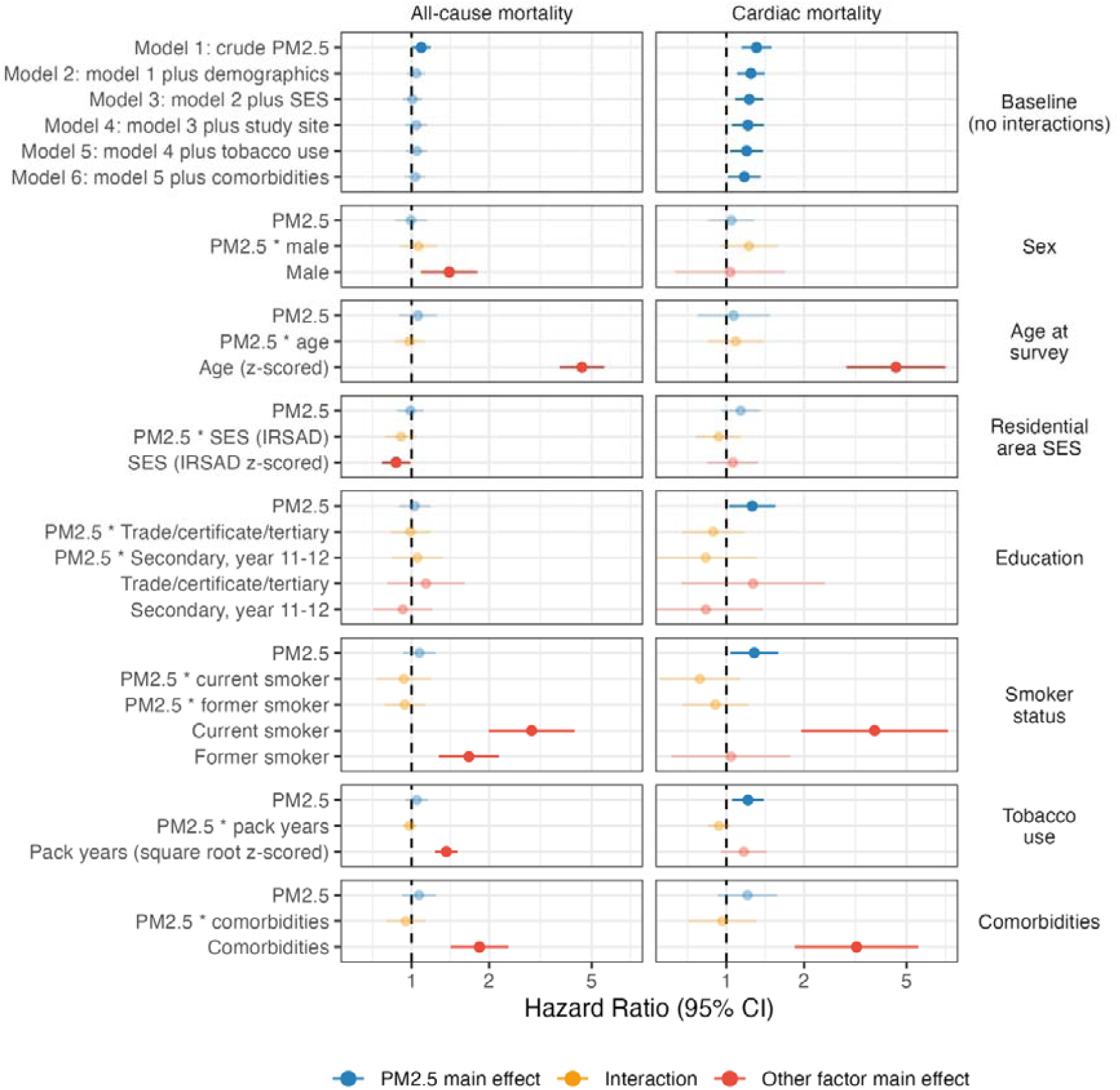
Forest plot of PM_2.5_ (per 10µg/m^3^ increase) effects on mortality, from crude to adjusted models, and moderated effects in fully-adjusted models.

In the main effects models 1-6, which do not include interaction terms, PM_2.5_ exposure had no detectable effect on all-cause mortality except in the Model 1, which only evaluated the crude (unadjusted) association. However, main effects unadjusted and adjusted models of cardiac-related mortality found that increasing PM_2.5_ exposure was associated with increasing mortality in a dose-response relationship. The association attenuated with each additional adjustment in models without interaction terms, with the smallest effect being an HR of 1.17 (95%CI 1.01-1.36) per 10µg/m^3^ of fire-related PM_2.5_ (fully adjusted model 6).

None of the tested potential moderators emerged as either protective against or an exacerbator of PM_2.5_ effects.

## 4 Discussion

Nine years after the Hazelwood coal mine fire of 2014, this study explored the rates of mortality from all-causes and cardiac causes amongst people living in the vicinity at the time. Some were from Morwell, the town that was covered with dense smoke from the fire, and the remainder from Sale, a similar town minimally affected by smoke. In these analyses, rates of mortality from all-causes were not found to be elevated, except in the crude model. This meant that, with this study size and design, mine fire smoke exposure had no detectable association with all-cause mortality in exposed cohort members. However, cardiac mortality rates were found to be increased in a dose response relationship with PM_2.5_ exposure at the time of the coal mine fire. As expected, cardiac mortality was also associated with increasing age, cigarette smoking and comorbidities. However, the effects of PM_2.5_ exposure on cardiac mortality were robust to sequential adjustment for demographic factors, socioeconomic status, study site, tobacco use, and comorbidities, although attenuated somewhat with each adjustment. None of these factors showed evidence of moderating the impact of PM_2.5_ exposure on cardiac mortality.

With regard to chronic exposure to the effects of air pollution, data from the Global Burden of Disease Study suggested that an estimated 6.7 million deaths in 2019 could be attributed to the effects of air pollution (31). The authors found that after tobacco, elevated systolic blood pressure and dietary risk, pollution was the fourth most important risk factor for mortality globally, more important than obesity, cholesterol, physical inactivity or alcohol excess. These effects could be attributed to both indoor air pollution (predominantly from use of biomass for cooking or kerosene lamps for lighting) (32) and outdoor air pollution (33). Of the excess deaths attributable to pollution, an estimated 50% were due to cardiovascular causes (31), accounting for almost 20% of all global deaths from cardiovascular disease. Whilst these researchers found evidence of an encouraging reduction in the exposure to household air pollution through socio-economic development, they found evidence of “concerning” increases in exposure to ambient particulate matter pollution in excess of 0.5% annually (31).

Air pollution not only exacerbates the course of cardiovascular diseases, but also increases the onset through combinations of mechanisms which may include: local lung inflammatory responses triggered by particulate matter; penetration of particulate matter deep into the lungs and therefore the bloodstream causing direct activation of systemic inflammation; oxidative stress; elevated stress hormones and insulin resistance; imbalance of autonomic and sympathetic nervous system function; impacts on the microbiome of the gut which may increase inflammation, atherosclerosis and metabolic syndrome; and direct damage to respiratory mucosa which leads to increased permeability for cardiotropic micro-organisms (34).

Although air pollution is a complex, dynamic mixture of gases and particles from a diversity of sources, three common pollutants are most commonly the focus of research, measurement and communication: ozone, nitrogen dioxide and particulate matter (35). Of these, the most consistent evidence about effects on cardiovascular disease has been found to date for particulate matter, which has established impacts on ischemic heart disease and stroke. Not only have these impacts been shown on short-term mortality (36) but also longer-term impacts on incidence and mortality of cardiovascular disease have been found (37).

Until now, there has been limited research about the mortality caused by pollution from coal mine fires (2). However, Liu and colleagues undertook a systematic review of the health effects of a different types of acute exposure to outdoor air pollution, namely that caused by forest fires, wildfires and peat fires (38). Including studies published between 1986-2014, the authors identified 13 studies of mortality after wildfire smoke exposure, among which nine showed increased rates of all-cause mortality. However, this review highlighted that 10 of 13 studies included accidental deaths related to the wildfires in their all-cause mortality rates (5,6,13); one of which (13) found no increased all-cause mortality rate after short-term exposure to wildfires.

Unfortunately, the reviewers found no studies of cardiac mortality specifically, but identified 14 studies which considered cardiac morbidity following acute exposure to wildfire smoke, assessed by presentations or admissions with cardiac arrest or symptoms of a cardiovascular disease after the exposure occurred (38). Interestingly, they reported some geographic variation such that five out of six studies examining exposure to wildfire smoke in USA were associated with increased hospital admissions for cardiovascular diseases (including cardiac arrest and chest pain). In contrast, seven studies from Australia and Canada found no impact on cardiovascular morbidity, measured similarly. Only one other study in the city of Porto, Portugal, found a significant increase in admissions for: hypertensive disease; ischaemic heart disease; and other cardiac diseases, including heart failure over a 3-month spate of summer forest fires in 2005 (39). The reviewers did not put forward a hypothesis to explain the apparent geographic variation although they pointed out that USA had much higher rates of cardiovascular diseases at the time than many of the other areas studied (38).

Although our findings are concordant with the growing body of literature suggesting a long-term impact of exposure to airborne particulate matter in polluted air, they were unexpected given that the participants in this cohort study could only be recruited 2 years after the coal mine fire. We have already shown that there was an excess of deaths from cardiovascular disease associated with PM_2.5_ exposure within 6 months of the mine fire (14). Consequently, the individuals who died during the fire or immediately afterwards could not be recruited into this cohort study. This has likely introduced survivor bias into the cohort where these early deaths, which could also be attributed to the mine fire, have acted to deplete the most susceptible members of the community prior to recruitment (40). For this reason, we suggest that the estimated excess cardiac mortality reported here is likely to be under-estimated.

This study has some strengths: the analyses were based on an established cohort for whom we had collected considerable information about individual risk factors, supplemented with diaries about their movements during the mine fire period. This enabled individual-level estimation of their PM_2.5_ exposure. Furthermore, the data about mortality have been extracted up to nine years after the mine fire from administrative databases, providing objective and comprehensive data that are known to be accurate and take account of deaths occurring outside the State of Victoria.

However, the findings must be considered alongside some limitations. Firstly, although the total follow-up period has been nine years following the mine fire, the number of deaths was still relatively small, limiting the statistical power of the current analyses. For this reason, more follow-up is still needed to know for sure whether deaths from all-causes have increased, and to precisely estimate the additional cardiac deaths caused by exposure to the coal mine fire. Secondly, the follow-up of this particular cohort for mortality took place after a number of deaths occurred during the mine fire and during the following 6 months. This has created a bias in those eligible to be recruited, so that the risk ratios provided are likely to be under-estimated.

In summary, this longitudinal analysis of mortality up to nine years after the Hazelwood Mine fire found no detectable increase in all-cause mortality, but an excess risk of deaths from cardiac causes, which appears to be attributable to the effects of individual-level PM exposure.

## Data Availability

All data produced in the present study are available upon reasonable request to the authors

https://doi.org/10.26180/28208279.v1

## Supplementary materials

**Table S1.** Associations between fire-related PM_2.5_ exposure and mortality in the Hazelwood Health Study Adult Cohort.

|  | All-cause mortality |  | Cardiac mortality |  |
| --- | --- | --- | --- | --- |
|  | HR (95% CI) | p-value | HR (95% CI) | p-value |
| **Baseline models (no interaction)** |  |  |  |  |
| Model 1: crude fire-related PM<sub>2.5</sub> exposure | **1.09 (1.00 - 1.19)** | **0.048** | **1.31 (1.14 - 1.50)** | **< 0.001** |
| Model 2: model 1 plus demographics | 1.04 (0.96 - 1.13) | 0.313 | **1.24 (1.10 - 1.41)** | **0.001** |
| Model 3: model 2 plus SES | 1.01 (0.93 - 1.10) | 0.837 | **1.23 (1.08 - 1.39)** | **0.002** |
| Model 4: model 3 plus study site | 1.04 (0.95 - 1.15) | 0.375 | **1.21 (1.05 - 1.40)** | **0.009** |
| Model 5: model 4 plus tobacco use | 1.05 (0.95 - 1.15) | 0.334 | **1.20 (1.03 - 1.39)** | **0.016** |
| Model 6: model 5 plus comorbidities | 1.03 (0.94 - 1.14) | 0.503 | **1.17 (1.01 - 1.36)** | **0.033** |
| **Age model** |  |  |  |  |
| Fire-related PM<sub>2.5</sub> exposure | 1.06 (0.89 - 1.25) | 0.509 | 1.07 (0.77 - 1.47) | 0.701 |
| PM<sub>2.5</sub> * age at Survey (z-score) | 0.98 (0.86 - 1.12) | 0.776 | 1.09 (0.84 - 1.39) | 0.521 |
| Age at Survey (z-score) | **4.58 (3.75 - 5.60)** | **<0.001** | 4.55 (2.92 - 7.08) | **<0.001** |
| **Sex model** |  |  |  |  |
| Fire-related PM<sub>2.5</sub> exposure | 1.00 (0.86 - 1.15) | 0.953 | 1.04 (0.85 - 1.28) | 0.676 |
| PM<sub>2.5</sub> * male | 1.06 (0.90 - 1.26) | 0.476 | 1.22 (0.94 - 1.59) | 0.133 |
| Male | **1.40 (1.09 - 1.81)** | **0.009** | 1.03 (0.63 - 1.69) | 0.896 |
| **Comorbidities model** |  |  |  |  |
| Fire-related PM<sub>2.5</sub> exposure | 1.07 (0.92 - 1.24) | 0.385 | 1.21 (0.93 - 1.57) | 0.165 |
| PM<sub>2.5</sub> * comorbidities | 0.95 (0.80 - 1.13) | 0.576 | 0.96 (0.71 - 1.31) | 0.815 |
| Any comorbidities<sup>†</sup> | 1.84 (1.42 - 2.38) | **<0.001** | 3.20 (1.84 - 5.55) | **<0.001** |
| **Socioeconomic area (Index of Relative Socioeconomic Advantage and Disadvantage 2016)** |  |  |  |  |
| Fire-related PM<sub>2.5</sub> exposure | 0.99 (0.88 - 1.11) | 0.875 | 1.14 (0.95 - 1.35) | 0.155 |
| PM<sub>2.5</sub> * IRSAD score (z-score) | 0.91 (0.79 - 1.05) | 0.200 | 0.93 (0.76 - 1.14) | 0.501 |
| IRSAD score (z-score) | **0.87 (0.77 - 0.99)** | **0.036** | 1.06 (0.85 - 1.33) | 0.619 |
| **Educational attainment (ref: secondary up to year 10)** |  |  |  |  |
| Fire-related PM<sub>2.5</sub> exposure | 1.03 (0.89 - 1.18) | 0.697 | **1.26 (1.02 - 1.55)** | **0.029** |
| PM<sub>2.5</sub> * certificate/diploma/tertiary | 1.05 (0.84 - 1.33) | 0.652 | 0.83 (0.53 - 1.31) | 0.428 |
| PM<sub>2.5</sub> * Secondary year 11-12 | 0.99 (0.82 - 1.19) | 0.915 | 0.89 (0.67 - 1.17) | 0.406 |
| Certificate/diploma/tertiary | 1.14 (0.80 - 1.61) | 0.464 | 1.27 (0.67 - 2.41) | 0.466 |
| Secondary year 11-12 | 0.93 (0.71 - 1.20) | 0.564 | 0.83 (0.50 - 1.38) | 0.478 |
| **Smoker status at survey** |  |  |  |  |
| Fire-related PM<sub>2.5</sub> exposure | 1.07 (0.93 - 1.24) | 0.340 | 1.28 (1.04 - 1.59) | 0.022 |
| PM<sub>2.5</sub> * former smoker | 0.94 (0.79 - 1.13) | 0.526 | 0.91 (0.67 - 1.22) | 0.514 |
| PM<sub>2.5</sub> * current smoker | 0.93 (0.73 - 1.19) | 0.585 | 0.79 (0.55 - 1.13) | 0.193 |
| Former smoker | 1.67 (1.28 - 2.18) | **<0.001** | 1.04 (0.61 - 1.77) | 0.882 |
| Current smoker | 2.92 (1.99 - 4.29) | **<0.001** | 3.76 (1.95 - 7.23) | **< 0.001** |
| **Cigarette pack-years at survey** |  |  |  |  |
| Fire-related PM<sub>2.5</sub> exposure | 1.05 (0.95 - 1.16) | 0.367 | **1.21 (1.05 - 1.40)** | **0.008** |
| PM<sub>2.5</sub> * pack-years (square root) | 0.98 (0.92 - 1.05) | 0.595 | 0.94 (0.85 - 1.04) | 0.209 |
| Pack-years (square-root) | 1.37 (1.23 - 1.51) | **<0.001** | 1.17 (0.95 - 1.43) | 0.137 |

| **Educational attainment (ref: secondary up to year 10)** |  |  |  |  |
| --- | --- | --- | --- | --- |
| Fire-related PM<sub>2.5</sub> exposure | 1.03 (0.89 - 1.18) | 0.697 | **1.26 (1.02 - 1.55)** | **0.029** |
| PM<sub>2.5</sub> * certificate/diploma/tertiary | 1.05 (0.84 - 1.33) | 0.652 | 0.83 (0.53 - 1.31) | 0.428 |
| PM<sub>2.5</sub> * Secondary year 11-12 | 0.99 (0.82 - 1.19) | 0.915 | 0.89 (0.67 - 1.17) | 0.406 |
| Certificate/diploma/tertiary | 1.14 (0.80 - 1.61) | 0.464 | 1.27 (0.67 - 2.41) | 0.466 |
| Secondary year 11-12 | 0.93 (0.71 - 1.20) | 0.564 | 0.83 (0.50 - 1.38) | 0.478 |

**† Self-reported pre-fire conditions including angina, myocardial infarction, heart failure, stroke, COPD, cancer, and diabetes; SES includes socioeconomic area (IRSAD) and educational attainment; \* indicates interactions (linear combination)**

**Figure S1.**
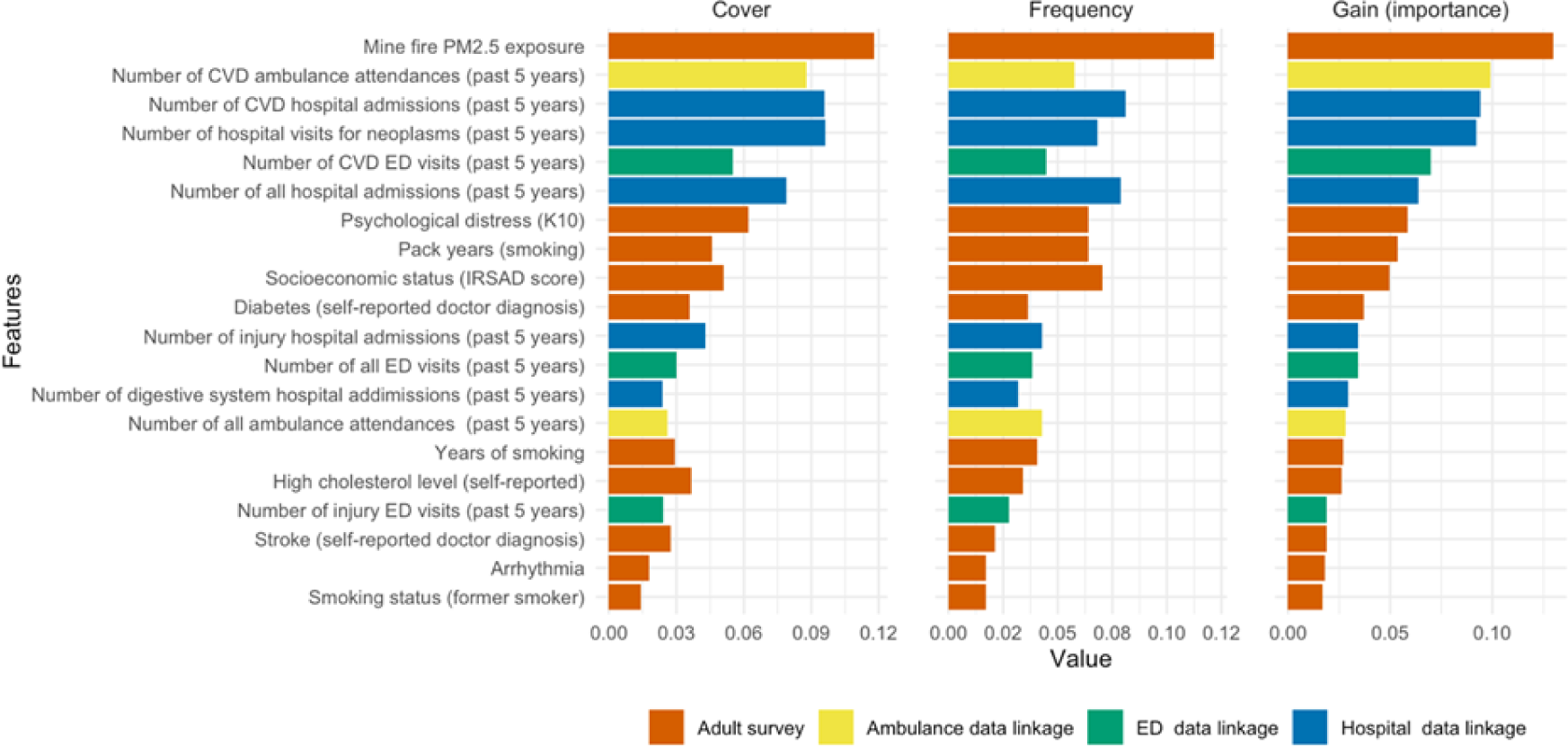
Variable importance plot of the top 20 factors in the prediction model for cardiac mortality among all mortality data. Notes. Cover: relative quantity of observations that a feature splits; Frequency: the number of times a feature is used to split data across all trees in the model; Gain: the improvement in accuracy (or reduction in loss) brought by a feature when it is used to split the data in the decision tree; ED: hospital emergency department; CVD: cardiovascular disease.

